# Are viral loads in the febrile phase a predictive factor of dengue disease severity?

**DOI:** 10.1101/2023.07.31.23293412

**Authors:** Shashika Dayarathna, Heshan Kuruppu, Tehani Silva, Laksiri Gomes, N.L. Ajantha Shyamali, Chandima Jeewandara, Dinuka Ariyaratne, Shyrar Tanussiya Ramu, Ananda Wijewickrama, Graham S. Ogg, Gathsaurie Neelika Malavige

## Abstract

**Background:** As many studies have shown conflicting results regarding the extent of viraemia and clinical disease severity, we sought to investigate if viraemia during early dengue illness is associated with subsequent clinical disease severity.

**Methodology/Principal Findings:** 424 adult patients, in whom the dengue virus (DENV) serotype could be identified, who presented within the first 4 days of illness, were recruited from a tertiary care hospital from Sri Lanka from September 2016 to September 2022 following informed written consent. To characterize subsequent clinical disease severity, all patients were followed throughout their illness daily and disease severity classified according to WHO 1997 and 2009 disease classification.

315 patients had DF, 109 progressed to develop DHF and of those 17 developed shock (DSS). Although the viral loads were higher in the febrile phase in patients who progressed to develop DHF than in patients with DF this was not significant (p=0.15). Significant differences were observed in viral loads in patients infected with different DENV serotypes (p=0.0001), with patients infected with DENV2 having the lowest viral loads and the highest viral loads in DENV1. Although those infected with DENV2 had lower viral loads, infection with DENV2 was significantly associated with a higher risk of developing DHF (p=0.016, Odds ratio 1.8; 95% CI 1.116 to 2.905). Based on the WHO 2009 disease classification, 268 had dengue with warning signs (DWW), 139 dengue without warning signs (DWoWS), and 17 had severe dengue (SD). No significant difference was observed in the viral loads between those with SD, DWW and DWoWS (p=0.34).

**Conclusions/Significance:** Viral loads in the febrile phase do not appear to significantly associate with subsequent clinical disease severity in a large Sri Lankan cohort.

**Author summary:** As many studies have shown conflicting results regarding the extent of viraemia and clinical disease severity, we sought to investigate if viraemia during early dengue illness was associated with subsequent clinical disease severity. We assessed the viral loads and subsequent clinical disease severity in 424 patients, during early illness, to determine if viral loads associate with disease subsequent disease severity. Although the viral loads were higher in early illness in patients who progressed to develop dengue haemorrhagic fever (DHF) than in patients with dengue fever, this was not significant. Significant differences were observed in viral loads in patients infected with different DENV serotypes, with patients infected with DENV2 having the lowest viral loads and the highest viral loads in DENV1. Although those infected with DENV2 had lower viral loads, infection with DENV2 was significantly associated with a higher risk of developing DHF. Therefore, viral loads in early illness, do not appear to strongly associate with subsequent clinical disease severity in this Sri Lankan cohort.

## Introduction

Dengue, which was named as one of the top ten threats to global health by the WHO is the most rapidly increasing mosquito borne viral infection [1]. As it is a climate sensitive infection, due to the rise in global temperatures, it is predicted that burden of infection due to dengue is likely to further increase, resulting in potential overwhelming of the health-care systems in further areas and countries [2]. As there is no effective treatment for dengue, all patients with a suspected dengue infection are closely monitored for early detection of complications for timely interventions in the form of meticulous fluid management [3].

While most dengue infections result in asymptomatic or mild illness, some individuals develop vascular leakage resulting in pleural effusions, ascites, shock and other complications such as bleeding and organ dysfunction [3]. The reasons for occurrence of vascular leakage and other complications in some individuals are unknown, but a secondary dengue infection, pregnancy, presence of comorbidities such as diabetes, obesity and chronic kidney disease are known risk factors [4–6]. A dysfunctional innate immune response that results in an impaired antiviral immunity, with an enhanced proinflammatory response, presence of poor quality non-neutralizing antibodies and a sub-optimum T cell response are known to contribute to severe disease [7, 8].

Although severe clinical disease manifestations occur due to an aberrant and suboptimum immune response, the dengue virus (DENV) is known to lead to induction of many inflammatory mediators and therefore, directly contributes to disease pathogenesis [9]. Therefore, targeting the DENV and inhibiting viral replication has been one of the main strategies for developing a treatment for dengue [10]. However, many studies have shown conflicting results regarding the extent of viraemia and clinical disease severity. In a recent very large study from Vietnam, it was shown that higher plasma viraemia increased the risk of severe disease, hospitalization and plasma leakage, irrespective of the infecting serotype and the immune status [11]. However, they also show that although the viral loads were highest for DENV1 and lowest with infections with DENV2, infection with DENV2 was associated with a higher risk of developing severe dengue [11]. Another study from Vietnam showed that viral loads during early illness was significantly less in those with secondary dengue infection, although secondary dengue is an important risk factor for severe dengue [12]. A study from Indonesia showed that DENV3 was associated with a higher risk of plasma leakage compared to other serotypes and that there was a trend towards higher viraemia in those who develop severe dengue [13]. Studies from Colombia and India showed that the viral loads had no relationship with clinical disease severity [14, 15].

The contradictory findings in different studies could be due to different viral dynamics based on timing, different genotypes of the virus, differences in host responses based on genetic predisposition and ethnicities. As many antiviral trials for dengue failed to show efficacy so far, and as many trials are underway, it would be important to understand if viral loads during early illness associate with subsequent disease severity in different countries and in different populations. In this study, we have characterized the viral loads during early illness in patients presenting to a large tertiary care hospital in Sri Lanka, over a period of 6 years, to understand the relationship between viral loads, different DENV serotypes and clinical disease severity.

## Methodology

### Patients with acute dengue infection

Adult patients (n=502) with a suspected acute dengue infection were recruited from the National Institute of Infectious Diseases, Sri Lanka from September 2016 to September 2022 following informed written consent to the study. Blood samples was collected from each patient, during the first four days since onset of symptoms (days of illness ≤ 4 days), which was during the febrile phase. A second blood sample could only be collected from 161 patients at day 5 to 7 of illness for dengue antibody assays. Any patient who had features of plasma leakage at the time of recruitment, were excluded from the study. To characterize clinical disease severity, all patients were followed throughout their illness daily, and all clinical and laboratory features were recorded. If the patients showed a rise in the haematocrit, with subsequent reduction in platelet counts (these parameters were monitored on a daily basis in all patients), ultrasound scans were performed to detect pleural effusions and ascites.

### Ethics approval

Ethical approval was obtained from the Ethics Review Committee of the Faculty of Medical Sciences, University of Sri Jayewardenepura.

### Determining the DENV serotype and viral loads

Blood samples were centrifuged, and the obtained serum samples were aliquoted and stored at -80°C to avoid repeated freeze-thawing. Viral RNA was extracted from the serum using QIAmp Viral RNA Mini Kit (Qiagen, USA, Cat: 52906). cDNA transcription was performed with a high-capacity cDNA reverse transcription kit (Applied Biosystems, USA, cat: 4368814).

Oligonucleotide primers, dual labelled probes for DEN 1 to 4 serotypes (Life Technologies, India) and TaqMan Multiplex Master Mix (Applied Biosystems, USA, Cat: 4461881) were used for the multiplex quantitative real-time PCR in ABI 7500 real time PCR system (Applied Biosystems, USA) as previously described [16]. The standard curve was generated using four gBlock fragments with known copy numbers (Integrated DNA Technologies, USA).

### Dengue IgM and IgG antibody assays

Dengue antibody assays were performed in 161 patients, in whom a second sample could be collected between day 5 to 7 of illness, using a commercial capture-IgM and IgG Enzyme-Linked Immunosorbent Assay (ELISA) (Panbio, Australia). Results were interpreted according to the manufacturer’s instructions. According to the WHO 2011 criteria, patients with an IgM: IgG ratio of >1.2 were classified as having a primary dengue infection, while patients with IgM: IgG ratios <1.2 were categorized under secondary dengue infection [3].

### Statistical analysis

Statistical analysis was done using GraphPad Prism version 9.5.1 (Dotmatics, California, USA). As the data were not normally distributed, differences in the viral loads for different clinical disease severity and serotypes were compared using the Mann-Whitney U test (two tailed). The Kruskal-Wallis test was used to compare the viral loads between different serotypes, when more than one parameter was compared. Spearman rank order correlation coefficient was used to evaluate the correlation between variables including the association between viral loads and laboratory parameters.

## Results

### Patient characteristics

Of the 502 patients recruited, the DENV serotype was identified in 424 (84.5%) patients and were included in the analysis. In order to determine the relationship between viral loads in the febrile phase and subsequent clinical disease severity, patients were classified as having DF or DHF, according to the WHO 1997 criteria[3], and analysis was also carried out by classifying them as having dengue with warning signs (DWW), without warning signs (DWoWS) and with severe disease (SD) according to the WHO 2009 disease classification guidelines [17]. The clinical and laboratory characteristics of patients when classified as DF/DHF, and DWW, DWoWS and SD are shown in table 1 and 2. The mean age of those who were recruited to the study was 33.28 years (SD ± 13.73) and median age 29.5 years. 241 (56.8%) patients were in the 20- to 40-year-old age group. There was no correlation with the age and the viral loads during the febrile phase of illness (Spearman’s R=-0.07, p=0.146).

**Table 1:** Clinical and laboratory features of patients who were classified as having DHF and DF based on the WHO 1997 disease classification.

| **Clinical findings** | **DF (n=315)**<br>**N (%)** | **DHF (n=109)**<br>**N (%)** |
| --- | --- | --- |
| Fever | 267 (84.6) | 102 (93.6) |
| Arthralgia | 177 (56.2) | 78 (71.6) |
| Myalgia | 173 (54.9) | 72 (66.1) |
| Flushing | 22 (7.0) | 7 (6.4) |
| Rashes | 4 (1.3) | 2 (1.8) |
| Itching | 5 (1.6) | 3 (2.8) |
| Abdominal pain | 56 (17.8) | 43 (39.4) |
| Vomiting | 64 (20.3) | 32 (29.4) |
| Diarrhoea | 83 (26.3) | 41 (37.6) |
| Pleural effusion | 0 | 17 (15.6) |
| Ascites | 0 | 90 (82.6) |
| Hepatomegaly | 3 (1) | 2 (1.8) |
| Epistaxis | 0 | 2 (1.8) |
| Haematemesis | 0 | 0 |
| Melena | 2 (0.6) | 0 |
| Vaginal bleeding | 5 (1.6) | 5 (4.6) |
| **Lowest platelet count /mm<sup>3</sup>** |  |  |
| <20,000 | 10 (3.2) | 50 (45.9) |
| 20,000 to 50,000 | 57 (18.1) | 44 (40.4) |
| 50,000 to 100,000 | 115 (36.5) | 9 (8.3) |
| >100,000 | 131 (41.6) | 5 (4.6) |

**Table 1: Clinical and laboratory features of patients who were classified as having DHF** 420 **and DF based on the WHO 1997 disease classification**
| **Lowest WBC count cells/μl** |  |  |
| --- | --- | --- |
| <2000 | 33 (10.5) | 14 (12.8) |
| 2000 to 5000 | 241 (76.5) | 85 (78.0) |
| >5000 | 39 (12.4) | 9 (8.3) |

**Table 2:**
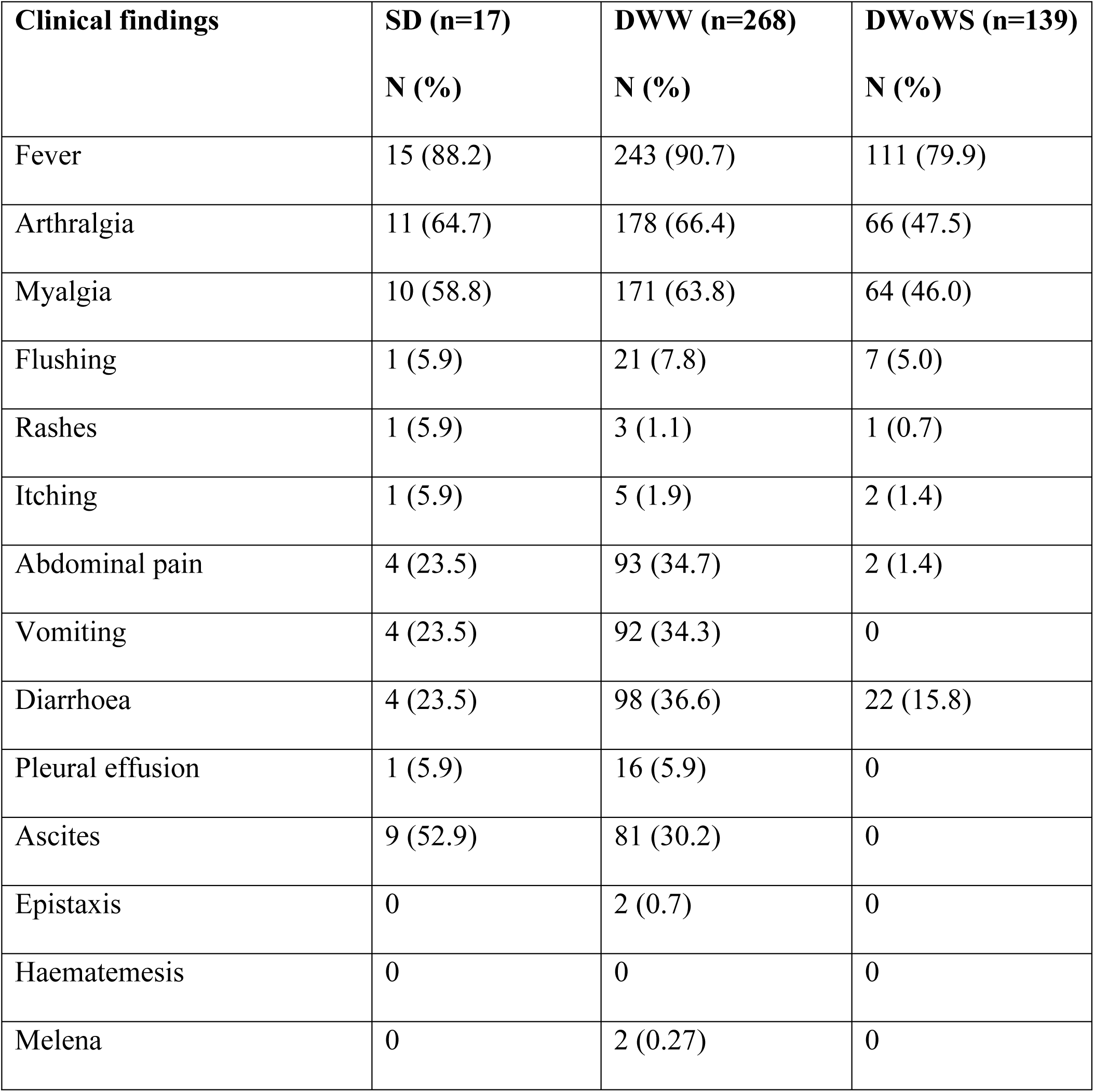
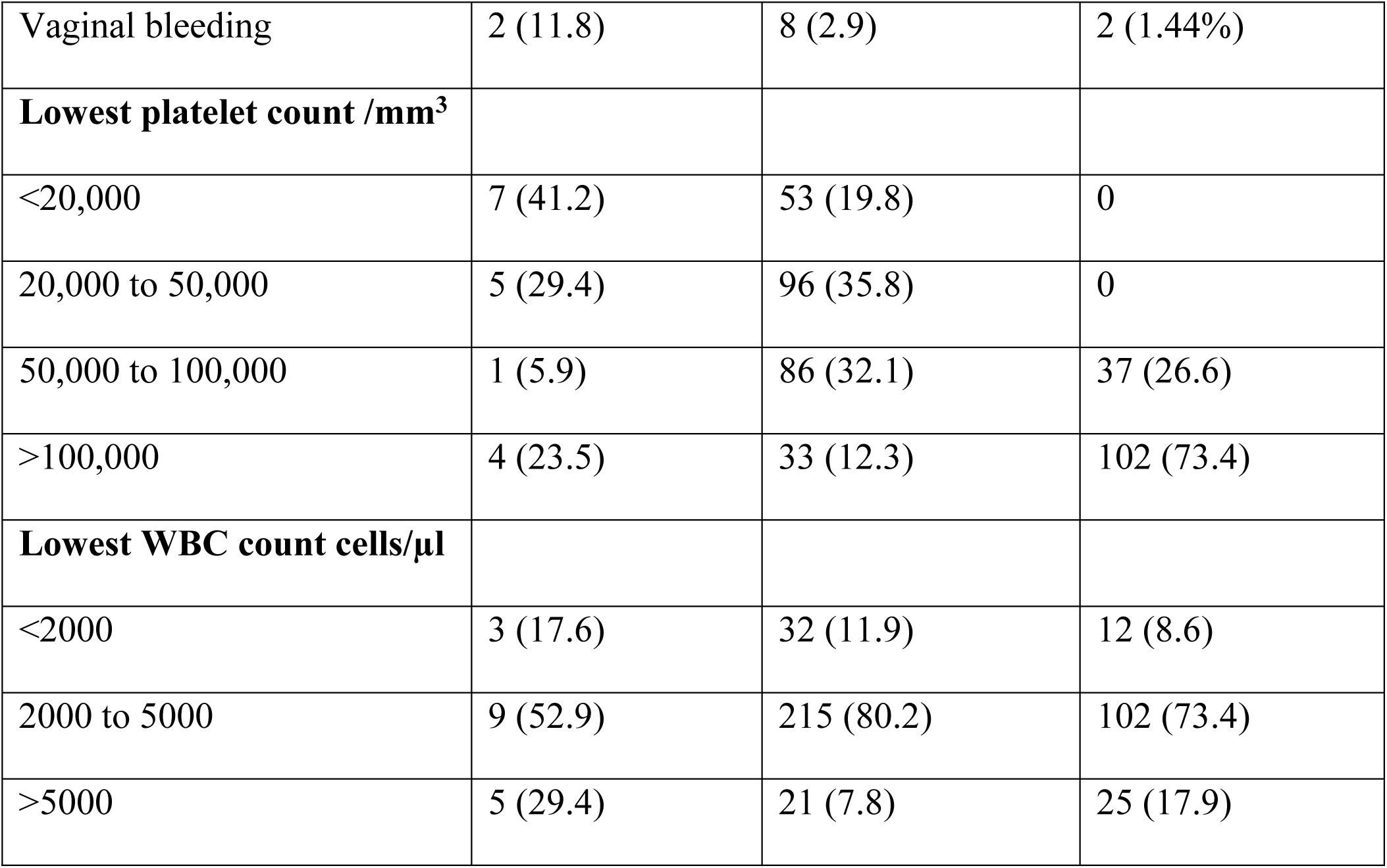
Clinical and laboratory features of patients who were classified as having severe dengue (SD), dengue with warning signs (DWW) and dengue without warning signs (DWoWS) according to the WHO 2009 disease classification.

According to the WHO 1997 guidelines, those who had features of plasma leakage (pleural effusions or ascites), along with platelet counts <100,000 cells/mm^3^ or a rise in haematocrit of >20% were classified as having DHF. Patients with DHF, who had a narrowing pulse pressure ≤20 were classified as having shock. Accordingly, 315 patients had DF, 109 progressed to develop DHF and of them 17 developed shock (DSS). Of those who had probable dengue (qPCR negative, but NS1 antigen test or dengue IgM/IgG rapid test positive), 52 had DF, 20 DHF and 6 patients DSS. Among the 424 qPCR positive patients, 273 (64.4%) were of males and 151 (35.6%) were of females, and 70 (25.6%) of males developed DHF vs 39 (25.8%) females.

Based on the WHO 2009 disease classification, 268 had DWW, 139 DWoWS, and 17 had SD. Of those who had probable dengue (qPCR negative, but NS1 antigen test or dengue IgM/IgG rapid test positive), 53 had DWW, 19 DWoWS and 6 had SD.

### Viral loads at febrile phase and relationship with clinical disease severity based on WHO 1997 disease classification

263 (62.0%) of patients were infected with DENV2, 97 (22.9%) with DENV1 and 61 (14.4%) with DENV3, while DENV4 was not detected in any of the patients. Three patients were found to be infected with two DENV serotypes: DENV1 and DENV2 (n=2) and DENV2 and DENV3 (n=1). Although the viral loads were higher in the febrile phase in patients who progressed to develop DHF (median 1.82×10^4^ copies/µl, IQR: 1.06×10^6^ copies/µl to 6.72×10^2^ copies/µl) than in patients with DF (median 7.85×10^3^ copies/µl, IQR: 7.80×10^5^ copies/µl to 1.41×10^2^ copies/µl), this was not significant (p=0.15) (Figure 1A). There were no significant differences in viral loads during the febrile phase in those who progressed to develop DSS vs DHF and DF (p=0.35).

**Figure 1.**
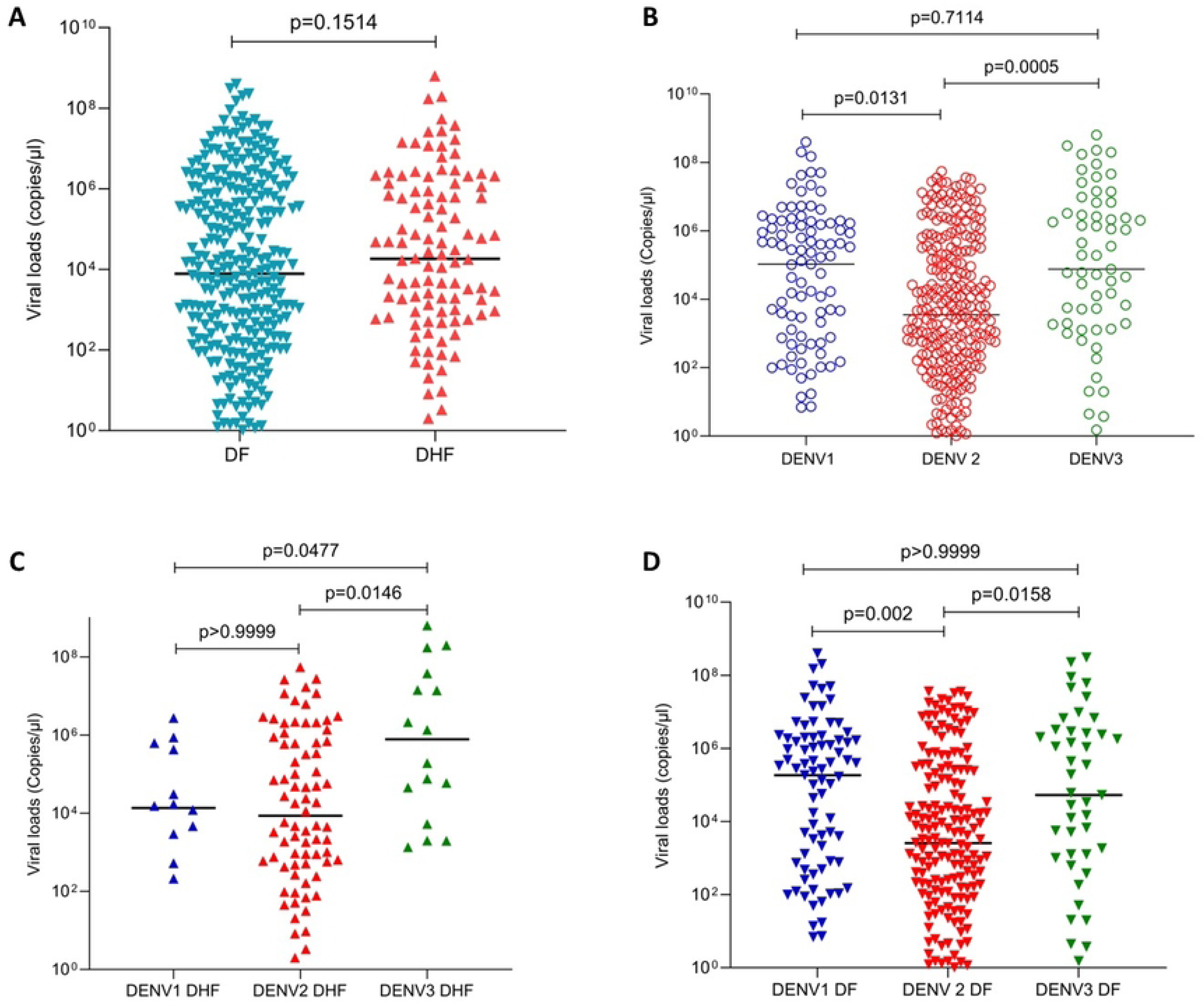
Viral loads in patients with varying clinical disease severity and infected with different DENV serotypes. Viral loads in serum samples were measured by qPCR in those with DF (n=313) and in those who progressed to DHF (n=108), during the febrile phase (A). Viral loads were compared in patients infected with different DENV virus serotypes, DENV1 (n=97), DENV2 (n=263) and DENV3 (n=61) (B) in those with DHF (C) and DF (D) infected with different virus serotypes. The Mann-Whitney test was used to compare the differences in viral loads between those with DF and DHF, while Kruskal-Wallis test was performed to compare the differences in viral loads in those infected different serotypes. All tests were two sided. Data are presented as median values +/- interquartile ranges as appropriate.

However, significant differences were observed in viral loads in patients infected with different DENV serotypes (p=0.0001) (Figure 1B). The lowest viral loads were detected in those infected with DENV2, while the highest viral loads were observed in those infected with DENV1. Although those infected with DENV2 had lower viral loads, infection with DENV2 was significantly associated with a higher risk of developing DHF (p=0.016, Odds ratio 1.8; 95% CI 1.116 to 2.905), when compared to being infected with other serotypes.

Significant differences were seen in the febrile phase in viral loads of patients infected with different viral serotypes, who subsequently progressed to develop DHF (p=0.013), (Figure 1C) or DF (p=0.0004), (Figure 1D). Although not significant, those who were infected with either DENV2 or DENV3 and subsequently progressed to DHF, had higher viral loads in the febrile phase than those who had DF. In contrast, in those infected with DENV1, those who progressed to DHF, had lower viral loads in the febrile phase than those who had DF.

Although a second blood sample was obtained in 161/502 initial cohort of patients, as the virus serotype could only be determined in 153/161 patients, we only included this cohort in the analysis of primary and secondary dengue. Accordingly, 49/153 (32.02%) had a primary dengue infection and 104/153 (67.97%) had a secondary dengue infection. There was no significant difference between the viral loads of those with primary and secondary dengue during early illness (p=0.6) (Figure 2). There was no difference between the viral loads in primary and secondary dengue in those infected with different DENV serotypes. However, the number of patients infected with DENV1 (n=32) and DENV3 (n=24) who were included in the analysis was too small for a meaningful analysis.

**Figure 2:**
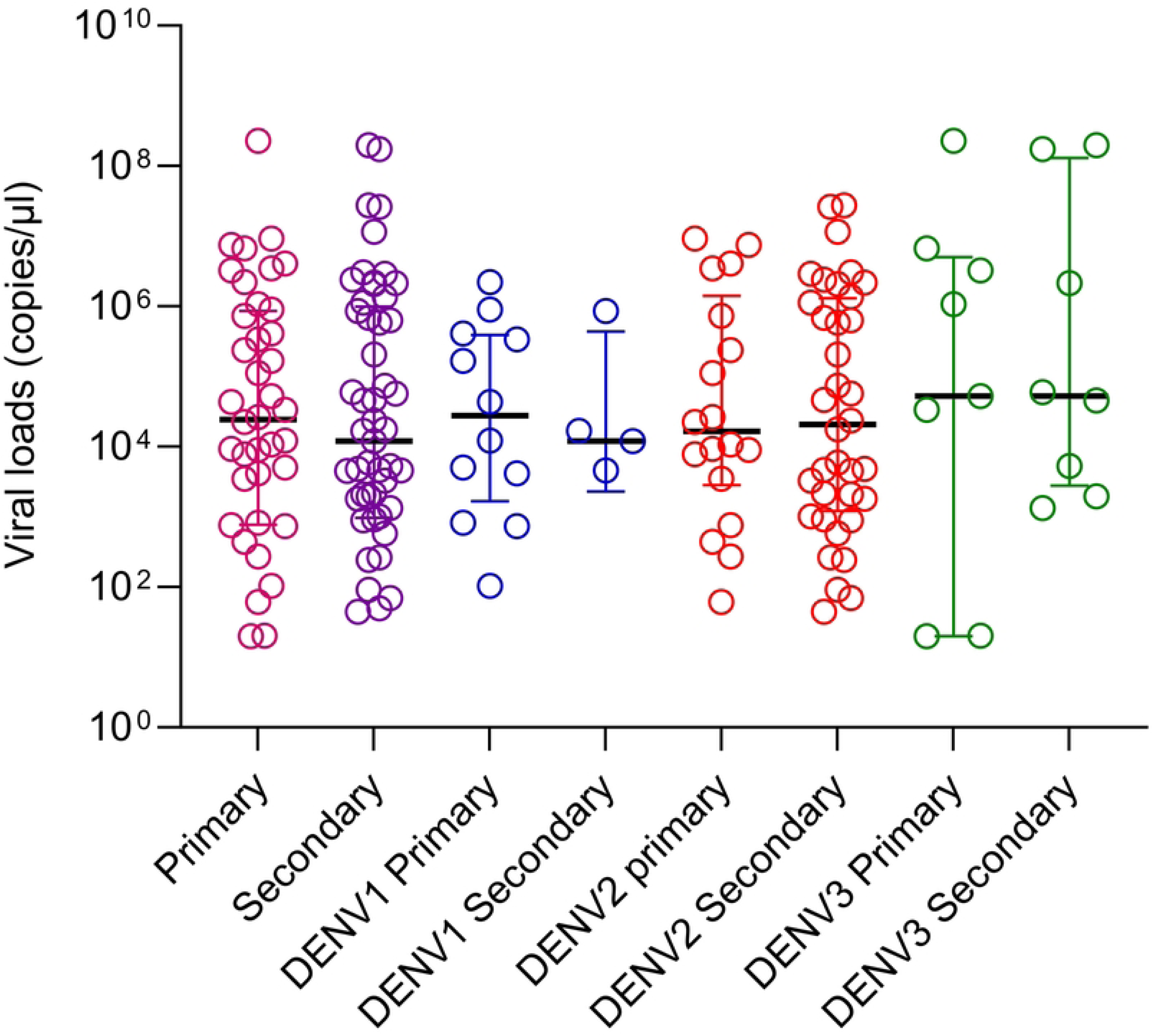
Viral loads in patients with primary and secondary dengue. Viral loads in serum samples were measured by qPCR in those primary (n=49) and secondary (n=104) dengue. They were also compared in those with primary and secondary dengue, infected with different DENV serotypes. The Mann-Whitney test was used to compare the differences in viral loads between those with primary and secondary dengue. All tests were two sided. Data are presented as median values +/- interquartile ranges as appropriate.

### Association of viral loads with laboratory parameters

The viral loads in the febrile phase inversely correlated with the lowest platelet counts of those who progressed to develop DHF (Spearman’s R= -0.24, p=0.01) (Figure 3A), but not with their lowest leucocyte counts (Spearman’s R= -0.06, p=0.56). No such association was seen in those with DF for platelet counts (Spearman’s R= 0.02, p=0.69) (Figure 3B) or leucocyte counts (Spearman’s R= -0.09, p=0.13). Liver enzymes (AST and ALT) could only be done in 179 patients, as these investigations were not routinely done throughout the years which the samples were collected. There was no correlation seen with the viral loads and ALT (Spearman’s R= -0.05, p=0.5) (Figure 3C) or AST (Spearman’s R= -0.006, p=0.9286) (Figure 3D). However, a significant difference was observed in AST (p=0.0005) and ALT (p=0.01) levels between those who developed DHF and DF.

**Figure 3.**
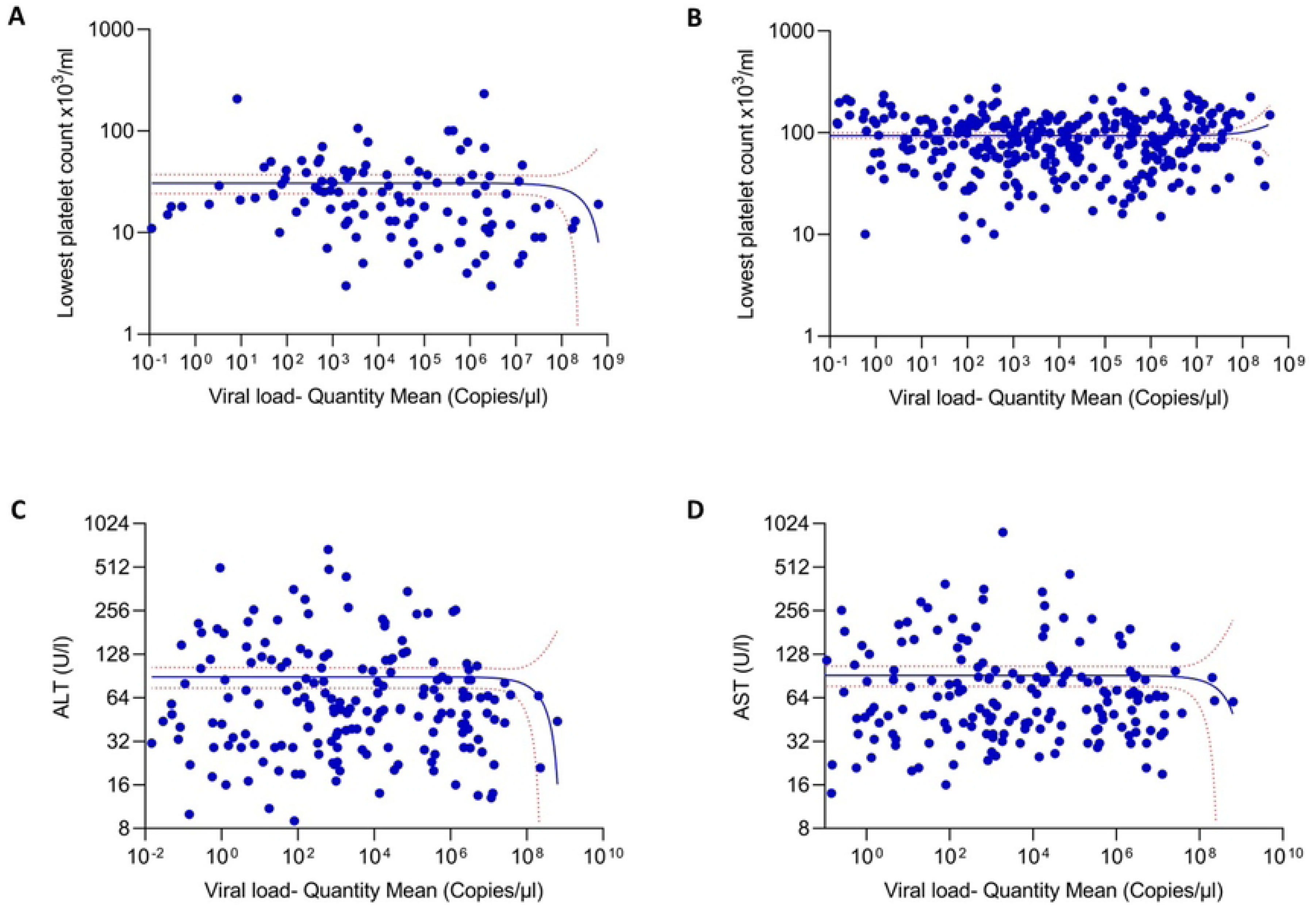
Association between the viral loads and the laboratory parameters in patients with varying severity of acute dengue. The correlation between the viral loads and the lowest platelet counts detected throughout the course of illness of the patients in DHF (n=108) (A), and DF (n=313) (B) were assessed. Data of 3 patients who had co-infection are not included in the analysis. Further, the viral loads and AST levels (C) and ALT levels (D) were measured in patients with DF and DHF patients (n=179). Spearman rank order correlation coefficient was used to evaluate the correlation between the viral loads and AST and ALT. All tests were two sided.

### Viral loads and relationship with clinical disease severity at the time of recruitment based on WHO 2009 criteria

No significant difference was observed in the viral loads between those with SD, DWW and DWoWS (p=0.34) (Figure 4A). However, significant differences were observed with the viral loads for different DENV serotypes, DWW (p=0.0001) (Figure 4B, C and D). Overall, the viral loads were lower with DENV2, compared to viral loads due to DENV1 and DENV2 in patients of all clinical disease categories.

**Figure 4.**
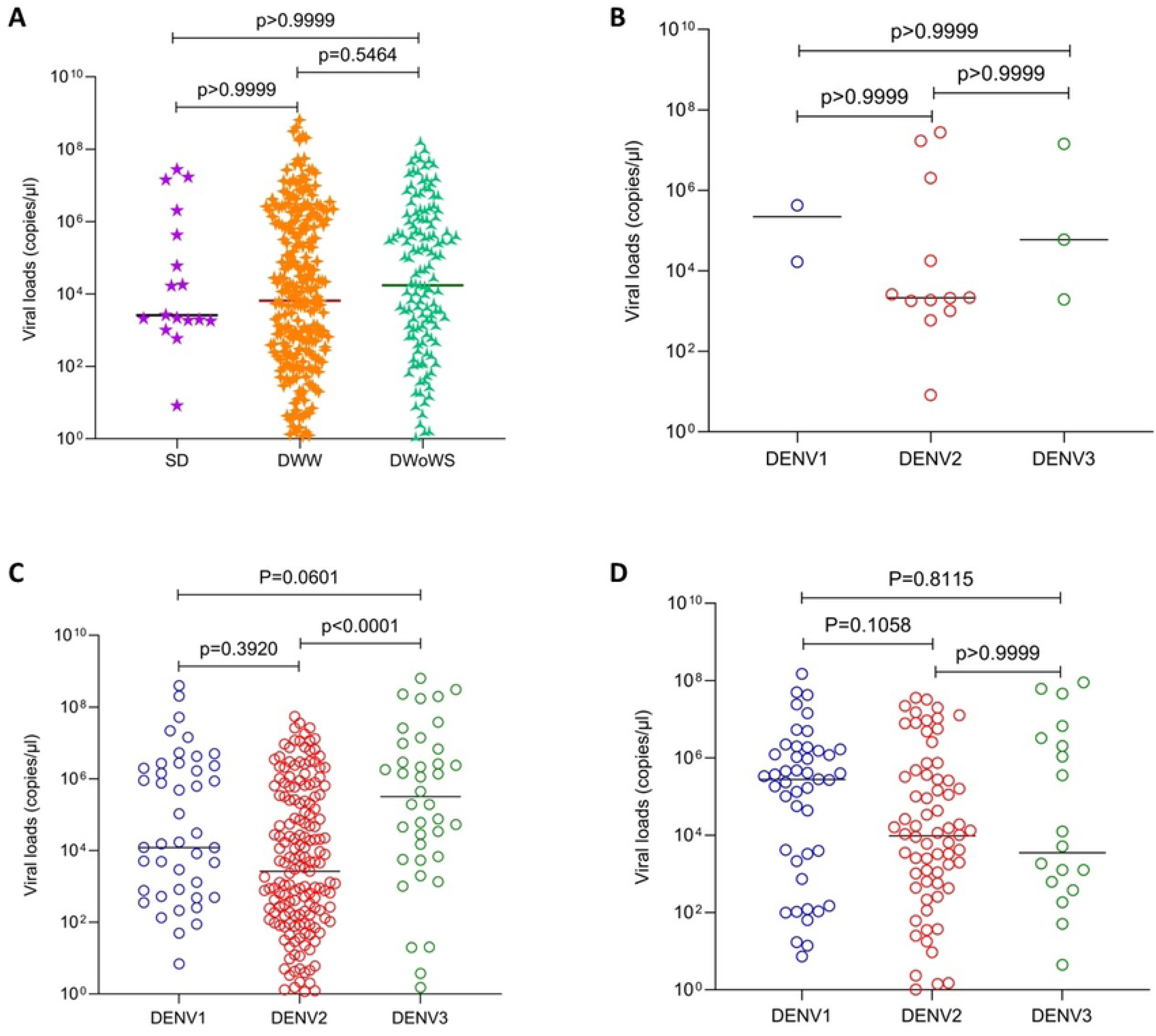
Viral loads in patients with varying clinical disease severity (classified according to the WHO 2009 disease classification) and infected with different DENV serotypes. Viral loads between patients with SD (n=17), DWW (n=266) and DWoWS (n=138) were compared (A). Data of 3 patients who had co-infection are not shown. Viral loads of different serotypes in patients with SD (n=17) (B), in those with DWW (n=266) (C) and in those with DWoWS (n=138) (D) were also compared. Kruskal-Wallis test was performed to compare the viral loads among the three groups of varying severity and the infected serotype. All tests were two sided. Data are presented as median values +/- interquartile ranges as appropriate and the p value is indicated.

## Discussion

In this study we have assessed the viral loads during early illness, during a period of six years in a large cohort of patients from Sri Lanka, who were followed up daily throughout the course of their illness, to determine their clinical disease severity. Using the WHO 1997 dengue disease classification we found that there was a trend towards viral loads being higher in those who progressed to DHF, although such a difference was not seen when the WHO 2009 dengue disease classification was used. As reported previously from a large study in Vietnam, and many other studies DENV2 was associated with lower viral loads, but with a higher risk of developing DHF [11, 18]. Interestingly, the viral loads were significantly higher in those infected with DENV3 in the febrile phase of those who progressed to DHF, compared to those infected with other serotypes, whereas the viral loads were highest for DENV1 in those who had DF. Higher viral loads for DENV1, followed by DENV3 has been shown in two previous studies from Vietnam [11, 12] further showing that the kinetics of viral loads significantly differ between the DENV serotypes during early illness. These differences in the viral loads and NS1 antigen levels for different DENV serotypes, have significant implications when evaluating point-of care diagnostics and antiviral drugs for dengue, as it would be important to assess them across different geographical regions and populations.

Although many studies do not show a statistically significant association with viral loads and clinical disease severity, there is a trend towards increased disease severity when infected with DENV2, which had the lowest viral loads of the DENV serotypes. It is possibly due to greater immune evasive capacity by DENV2 infection, as strains of DENV2 have shown an enhanced ability to inhibit type 1 interferon signaling compared to other DENV serotypes [19]. Apart from these variations seen between DENV serotypes and particular genotypes, there is a huge individual variation between viral loads. Our data along with many other studies show several log differences of viral loads for different DENV serotypes in patients with varying disease severity during the febrile phase. Many factors such as the initial viral inoculum, the incubation period, previous immunity, and presence of comorbid illnesses has shown to affect viral loads [20]. It was shown that the incubation period was shorter in those with secondary dengue infection compared to primary infection, supporting the role of antibody dependent enhancement in disease pathogenesis [20, 21].

Although we analyzed the differences in viral loads in the febrile phase in those with primary and secondary dengue, the sample size was small and only a few patients infected were infected with DENV1 and DENV3, were possible to include in analysis. However, our results were similar to the large study done in Vietnam comparing viral loads in relation to immune status, which showed no difference in early illness [11]. A previous study done in Vietnam and in Singapore for evaluation of the antiviral effect of celgosvir showed that the viral loads were not in fact different in the early phase, but those with secondary dengue, cleared the plasma viraemia earlier than those with primary dengue [12, 22]. Therefore, although patients with secondary dengue have shorter incubation periods and possibly higher viral infection in FcγR bearing cells, due to antibody dependent enhancement, this is not reflected in the viraemia seen in serum. This possibly suggests that the viral loads and viral kinetics seen in serum may not necessarily reflect the viraemia within immune cells and tissues.

In summary, although the degree of viraemia leads to severe disease, there is an important role played by the host-immune response to which could lead to resolution of infection or lead to immunopathogenesis. While use of antivirals would be an important treatment strategy for dengue, drugs that target certain immune mediators, such as leukotrienes, chymase, tryptase, platelet activating factor, growth factors, cytokines and other lipid mediators would be of potential benefit.

## Data Availability

Data is available within the manuscript and figures.

## Acknowledgements

We are grateful to the NIH, USA (grant number 5U01AI151788-02), Accelerating Higher Education Expansion and Development (AHEAD) Operation of the Ministry of Higher Education funded by the World Bank, and the UK Medical Research Council.

## References

1. Ten threats to global health in 2019 [Internet]. World Health Organization; 2019. Available from: https://www.who.int/news-room/spotlight/ten-threats-to-global-health-in-2019

2. Colon-Gonzalez FJ, Sewe MO, Tompkins AM, Sjodin H, Casallas A, Rocklov J, et al. Projecting the risk of mosquito-borne diseases in a warmer and more populated world: a multi-model, multi-scenario intercomparison modelling study. Lancet Planet Health. 2021;5(7):e404–e14. doi: 10.1016/S2542-5196(21)00132-7. PubMed PMID: 34245711; PubMed Central PMCID: PMCPMC8280459.

3. WHO, editor. Comprehensive guidelines for prevention and control of dengue fever and dengue haemorrhagic fever. SEARO, New Delhi, India: World Health Organization; 2011.

4. Malavige GN, Jeewandara C, Ogg GS. Dengue and COVID-19: two sides of the same coin. Journal of biomedical science. 2022;29(1):48. Epub 20220703. doi: 10.1186/s12929-022-00833-y. PubMed PMID: 35786403; PubMed Central PMCID: PMCPMC9251039.

5. Macias AE, Werneck GL, Castro R, Mascarenas C, Coudeville L, Morley D, et al. Mortality among Hospitalized Dengue Patients with Comorbidities in Mexico, Brazil, and Colombia. The American journal of tropical medicine and hygiene. 2021;105(1):102–9. Epub 20210510. doi: 10.4269/ajtmh.20-1163. PubMed PMID: 33970884; PubMed Central PMCID: PMCPMC8274750.

6. Sangkaew S, Ming D, Boonyasiri A, Honeyford K, Kalayanarooj S, Yacoub S, et al. Risk predictors of progression to severe disease during the febrile phase of dengue: a systematic review and meta-analysis. The Lancet infectious diseases. 2021;21(7):1014–26. Epub 20210225. doi: 10.1016/S1473-3099(20)30601-0. PubMed PMID: 33640077; PubMed Central PMCID: PMCPMC8240557.

7. Malavige GN, Jeewandara C, Ogg GS. Dysfunctional Innate Immune Responses and Severe Dengue. Front Cell Infect Microbiol. 2020;10:590004. doi: 10.3389/fcimb.2020.590004. PubMed PMID: 33194836; PubMed Central PMCID: PMCPMC7644808.

8. St John AL, Rathore APS. Adaptive immune responses to primary and secondary dengue virus infections. Nature reviews Immunology. 2019;19(4):218–30. doi: 10.1038/s41577-019-0123-x. PubMed PMID: 30679808.

9. Bhatt P, Sabeena SP, Varma M, Arunkumar G. Current Understanding of the Pathogenesis of Dengue Virus Infection. Curr Microbiol. 2021;78(1):17–32. Epub 20201124. doi: 10.1007/s00284-020-02284-w. PubMed PMID: 33231723; PubMed Central PMCID: PMCPMC7815537.

10. Troost B, Smit JM. Recent advances in antiviral drug development towards dengue virus. Current opinion in virology. 2020;43:9–21. Epub 20200811. doi: 10.1016/j.coviro.2020.07.009. PubMed PMID: 32795907.

11. Vuong NL, Quyen NTH, Tien NTH, Tuan NM, Kien DTH, Lam PK, et al. Higher Plasma Viremia in the Febrile Phase Is Associated With Adverse Dengue Outcomes Irrespective of Infecting Serotype or Host Immune Status: An Analysis of 5642 Vietnamese Cases. Clin Infect Dis. 2021;72(12):e1074–e83. doi: 10.1093/cid/ciaa1840. PubMed PMID: 33340040; PubMed Central PMCID: PMCPMC8204785.

12. Tricou V, Minh NN, Farrar J, Tran HT, Simmons CP. Kinetics of viremia and NS1 antigenemia are shaped by immune status and virus serotype in adults with dengue. PLoS neglected tropical diseases. 2011;5(9):e1309. Epub 2011/09/13. doi: 10.1371/journal.pntd.0001309. PubMed PMID: 21909448; PubMed Central PMCID: PMC3167785.

13. Nainggolan L, Dewi BE, Hakiki A, Pranata AJ, Sudiro TM, Martina B, et al. Association of viral kinetics, infection history, NS1 protein with plasma leakage among Indonesian dengue infected patients. PloS one. 2023;18(5):e0285087. Epub 20230502. doi: 10.1371/journal.pone.0285087. PubMed PMID: 37130105; PubMed Central PMCID: PMCPMC10153689.

14. Singla M, Kar M, Sethi T, Kabra SK, Lodha R, Chandele A, et al. Immune Response to Dengue Virus Infection in Pediatric Patients in New Delhi, India--Association of Viremia, Inflammatory Mediators and Monocytes with Disease Severity. PLoS neglected tropical diseases. 2016;10(3):e0004497. doi: 10.1371/journal.pntd.0004497. PubMed PMID: 26982706; PubMed Central PMCID: PMCPMC4794248.

15. Perdomo-Celis F, Salgado DM, Narvaez CF. Magnitude of viremia, antigenemia and infection of circulating monocytes in children with mild and severe dengue. Acta tropica. 2017;167:1–8. Epub 20161213. doi: 10.1016/j.actatropica.2016.12.011. PubMed PMID: 27986543.

16. Silva T, Jeewandara C, Gomes L, Gangani C, Mahapatuna SD, Pathmanathan T, et al. Urinary leukotrienes and histamine in patients with varying severity of acute dengue. PloS one. 2021;16(2):e0245926. Epub 20210205. doi: 10.1371/journal.pone.0245926. PubMed PMID: 33544746; PubMed Central PMCID: PMCPMC7864425.

17. WHO. Dengue guidelines, for diagnosis, treatment, prevention and control. 2009 21st April 2009. Report No.

18. Vicente CR, Herbinger KH, Froschl G, Malta Romano C, de Souza Areias Cabidelle A, Cerutti Junior C. Serotype influences on dengue severity: a cross-sectional study on 485 confirmed dengue cases in Vitoria, Brazil. BMC infectious diseases. 2016;16:320. Epub 20160708. doi: 10.1186/s12879-016-1668-y. PubMed PMID: 27393011; PubMed Central PMCID: PMCPMC4938938.

19. Medina FA, Torres-Malave G, Chase AJ, Santiago GA, Medina JF, Santiago LM, et al. Differences in type I interferon signaling antagonism by dengue viruses in human and non-human primate cell lines. PLoS neglected tropical diseases. 2015;9(3):e0003468. Epub 20150313. doi: 10.1371/journal.pntd.0003468. PubMed PMID: 25768016; PubMed Central PMCID: PMCPMC4359095.

20. Ben-Shachar R, Schmidler S, Koelle K. Drivers of Inter-individual Variation in Dengue Viral Load Dynamics. PLoS Comput Biol. 2016;12(11):e1005194. Epub 20161117. doi: 10.1371/journal.pcbi.1005194. PubMed PMID: 27855153; PubMed Central PMCID: PMCPMC5113863.

21. Riswari SF, Velies DS, Lukman N, Jaya UA, Djauhari H, Ma’roef CN, et al. Dengue incidence and length of viremia by RT-PCR in a prospective observational community contact cluster study from 2005-2009 in Indonesia. PLoS neglected tropical diseases. 2023;17(2):e0011104. Epub 20230206. doi: 10.1371/journal.pntd.0011104. PubMed PMID: 36745606; PubMed Central PMCID: PMCPMC9901748.

22. Sung C, Wei Y, Watanabe S, Lee HS, Khoo YM, Fan L, et al. Extended Evaluation of Virological, Immunological and Pharmacokinetic Endpoints of CELADEN: A Randomized, Placebo-Controlled Trial of Celgosivir in Dengue Fever Patients. PLoS neglected tropical diseases. 2016;10(8):e0004851. doi: 10.1371/journal.pntd.0004851. PubMed PMID: 27509020; PubMed Central PMCID: PMCPMC4980036.

